# Differential Diagnosis of Image Discovered Small Pulmonary Nodules (SPN): A Real-World Study

**DOI:** 10.1101/2025.07.02.25330729

**Authors:** Rongshan Zheng, Yilin Lai, Ji Huang, Nengluan Xu, Hongying Zhang, Jianfeng Xie, Hongru Li

## Abstract

**Objective:** The present investigation explored the disease spectrum of SPN (≤2cm) and attempted to establish the potential predictive models for non-benign small pulmonary nodules (NBSPN) identified from CT screening in a real-world perspective study.

**Methods:** A retrospective analysis was conducted on 6166 patients with SPN detected via chest CT scans at Fuzhou University Affiliated Provincial Hospital from January 2017 to September 2022. R language and SPSS software were used for data analyses.

**Results:** Of the 6,166 patients with SPN, 954 (15.47%) had their diagnosis confirmed pathologically. Among these nodules, NBSPN accounted for 88.7%. In NBSPN, the precursor glandular lesion (PGL) accounted for 14.18%, including 5.83% with atypical adenomatous hyperplasia (AAH) and 94.17% with adenocarcinoma in situ (AIS). Lung adenocarcinoma (LUAD) accounted for 85.82%, of which 32.07% were microinvasive adenocarcinoma (MIAD) and 67.93% invasive adenocarcinoma (IAD). Using the ROC model, none of the classical clinical factors had predictive value to the nature of SPN, including age, smoking, underlying conditions, CEA level, family history etc. Interestingly, the risk factors for NBSPN predicted by the ROC model include: female (OR: 1.842, 95%CI: 1.086-3.125, P=0.024), pure ground-glass opacities(pGGO) (OR: 5.243, 95%CI: 2.640-10.411, P<0.001), and part-solid (PS) (OR: 5.643, 95%CI: 1.970-16.167, P=0.001) in chest imaging, collectively delivering a highly significant AUC value at 0.748 in training set and 0.799 in validation set for distinguishing NBSPN from BSPN.

**Conclusion:** Conventional clinical data are not sufficient to differentiate NBSPN from BSPN. Factors with high predictive values for NBSPN include female, pGGO and PS.

## Introduction

According to the 2020 Global Cancer Database, lung cancer remains the leading cause of cancer-related deaths worldwide, accounting for approximately one-fifth (18%) of all cancer deaths.[1] Characterized by its high incidence and poor prognosis, lung cancer ranks as the top cause of cancer deaths. The five-year survival rate for advanced lung cancer is only around 16% . Early screening and diagnosis can elevate this rate to 70%-90%, highlighting the crucial importance of early detection and treatment, which has garnered significant attention in recent years.

Small pulmonary nodules (SPNs) are defined as focal, round-like lesions with a diameter of ≤ 2 cm on imaging, exhibiting higher density than the surrounding lung parenchyma and appearing as solid or subsolid nodules.[2][3][4] With increased health awareness and the widespread adoption of computed tomography (CT) technology, particularly advancements in high-resolution CT (HRCT) technology, pulmonary CT has gradually replaced chest X-rays as a common imaging examination in pulmonary health screenings. Consequently, there has been an increase in the detection of small to medium-sized pulmonary nodules (with an average diameter of 4 to 25 mm).[5] In recent years, research on pulmonary nodules has gradually increased, but there are still few reports using real-world data for discriminant analysis of different properties. The 2023 edition of Expert Consensus on Early Lung Cancer Diagnosis included meta-analysis and retrospective research data, and expert voting is used to recommend relevant risk factors.[6] Nevertheless, there is still limited discussion and research on distinguishing the nature of SPNs. The 2024 Chinese Expert Consensus on the Rational Diagnosis and Treatment of Pulmonary Nodules with a diameter of ≤ 2 cm mainly focus on follow-up and surgical procedures for SPNs,[7] but the discussion on the discrimination of pulmonary nodules is not deep enough. Therefore, it is necessary to conduct real-world data exploration to clinically predict the nature of SPNs, in order to improve the understanding of medical personnel and the public towards SPNs.

According to the Chinese Expert Consensus on the Diagnosis and Treatment of Pulmonary Nodules (2024),[8] it is recommended to lower the screening age for lung cancer to 40 years for individuals with any of the following risk factors: (1) smoking ≥ 400 pack-years (or 20 pack-years); (2) history of exposure to environmental or high-risk occupations (such as asbestos, beryllium, uranium, radon, etc.); (3) coexisting chronic obstructive pulmonary disease, diffuse pulmonary fibrosis, or a history of tuberculosis; (4) history of malignancy or family history of lung cancer, especially among first-degree relatives. However, the expert consensus mentioned that pulmonary nodules are those with a diameter of ≤ 3 cm. With the increasing detection rate of pulmonary nodules with a diameter of ≤ 2cm and the impact of modern society and environmental changes, further research is needed to ascertain whether the risk factors for malignant SPNs align with those traditionally identified for pulmonary nodules ≤ 3 cm.

Based on the amount of solid components on CT images, pulmonary nodules are classified into pure ground-glass nodules (pGGOs), partially solid nodules (PSNs), and solid nodules (SNs). Previous studies have shown that nodules with more solid components tend to grow rapidly, metastasize earlier, and are associated with a higher risk of malignancy, often leading to poorer prognosis.[9] In addition, many studies on CT imaging have suggested that spiculation, lobulation, vascular convergence, and pleural traction are related to the malignancy of lung cancer. Therefore, they help distinguish between benign and malignant nodules.[10] However, these imaging features are often not obvious in SPNs with a diameter of ≤ 2 cm, making it difficult for chest imaging to differentiate the nature of the nodules. At present, the diagnosis of pulmonary nodules is mainly based on the patient’s clinical data, combined with the results of chest CT screening results, and based on the proliferation characteristics of malignant tumors, the proliferation and imaging changes of nodule size are used for clinical follow-up and further discrimination. However, whether there is a better accuracy in distinguishing the nature of SPNs still needs indepth exploration.

Based on the existing problems mentioned above, this article retrospectively analyzes the clinical data of 6,166 cases of pulmonary nodules with a diameter ≤ 2 cm that were followed up at Fuzhou University Affiliated Provincial Hospital from January 1, 2017 to September 30, 2022. The clinical research big data platform was used to retrieve the previous medical information of the cases, and the follow-up date was January 17, 2023. By analyzing the clinical data, blood tumor markers, imaging features, pathological results, etc. of the cases, epidemiological data of pulmonary nodules with a diameter of ≤ 2 cm and high-risk factors of non-benign small pulmonary nodules (NBSPNs) with follow-up in our center were obtained. The predicted high-risk factors of NBSPNs were analyzed to explore the clinical and imaging characteristics of different types of malignant pulmonary nodules, in order to improve the ability to quickly and accurately distinguish the nature of pulmonary nodules and make corresponding diagnosis and treatment decisions in clinical practice.

## Materials and Methods

### 1. Study Population

Cases were retrieved from the Clinical Research Big Data Platform of Fuzhou University Affiliated Provincial Hospital for patients who underwent imaging examinations and were found to have SPNs ≤ 2 cm in diameter between January 1, 2017, and September 30, 2022, with at least one follow-up by January 17, 2023. The clinical data was screened according to the screening process (Figure 1).

**Figure 1.**
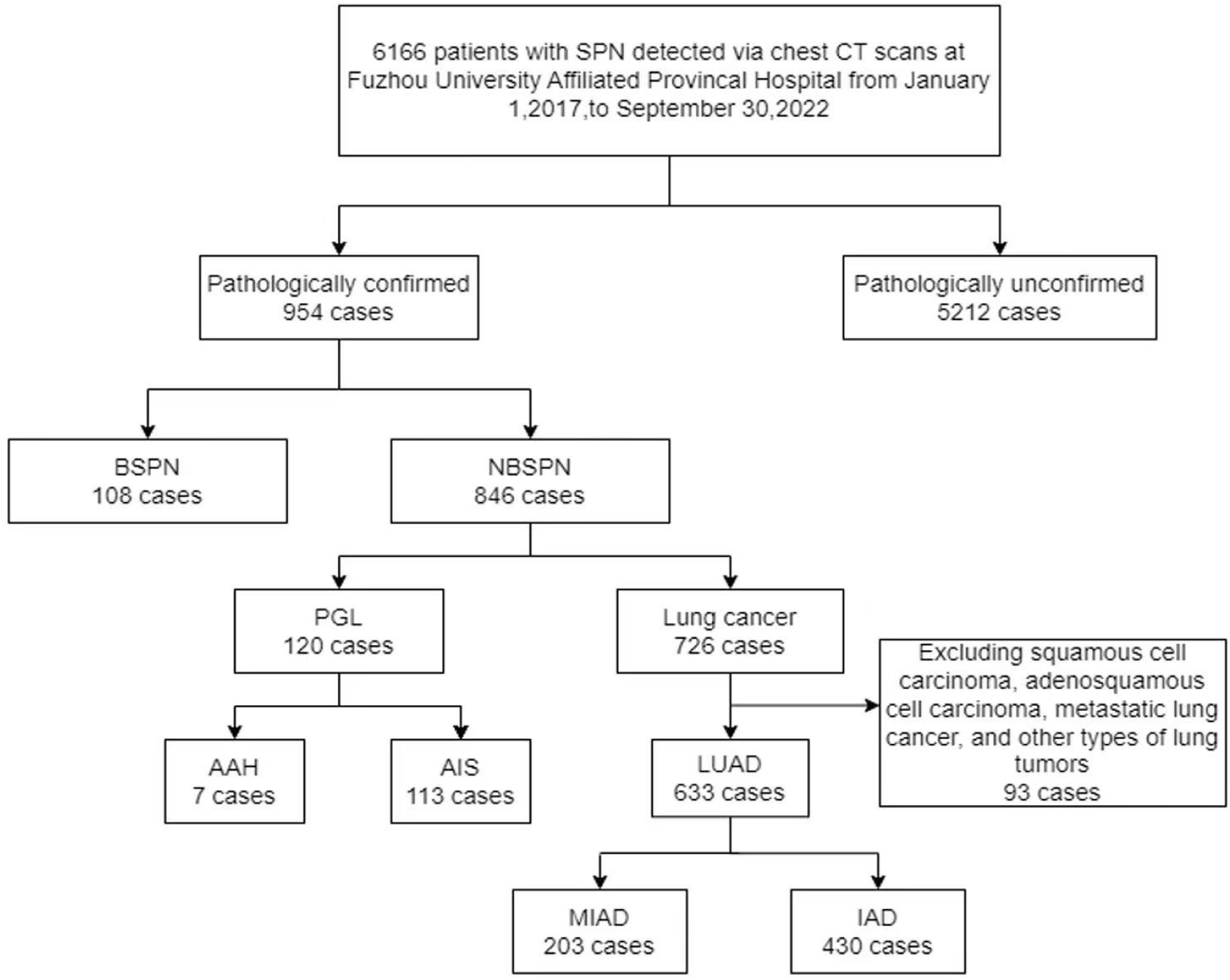
Screening Process Diagram.

Inclusion Criteria:

1. Confirmed presence of SPNs via chest/lung CT or whole-body PET/CT (Position Emission Tomography/Computed Tomography) ;
2. Meets the diagnostic criteria for SPNs, which are focal, solid or subsolid circular lesions with a diameter of less than or equal to 2 cm and increased density compared to lung parenchyma on imaging.

Exclusion Criteria:

1. Non-compiance with the diagnostic criteria for SPNs;
2. Incomplete clinical data.

### 2. Research methods

#### 2.1 Data Collection

This study adopted a retrospective research method, based on the medical record data collected by the clinical research big data platform system of Fuzhou University Affiliated Provincial Hospital, and included the clinical data of standard patients, including gender, age, medical history [chronic bronchitis, chronic obstructive pulmonary disease (COPD), interstitial pneumonia, tuberculosis, malignant tumors], family history of malignant tumors, smoking history, alcohol consumption history, tumor biomarkers [including carcinoembryonic antigen (CEA), cytokeratin fragment 19 (CYFRA21-1), squamous cell carcinoma antigen (SCC), neuron-specific enolase (NSE), pro-gastrin-releasing peptide (proGRP)], and pathological diagnosis results of SPNs. Collect the size and specific imaging features (pGGO, PSN, SN, spiculation, spinous processes, lobulation, vacuole sign, calcification, vascular convergence, pleural traction, blood vessel passage, air-bronchogram) of each assessable SPNs.

### 2.2 Data Grouping

A total of 954 cases of pulmonary nodules were identified through bronchoscopy or lung biopsy, as well as postoperative pathology, including 108 benign nodules (BSPNs) and 846 NBSPNs. BSPNs were classified as non-tumor and non-malignant nodules. NBSPNs were further divided into precursor glandular lesions (PGL) and lung cancer groups based on the interpretation of lung tumours in the WHO classification of thoracic tumours (5th edition).[11] PGL include atypical adenomatous hyperplasia (AAH) and adenocarcinoma in situ (AIS), while the lung cancer group includes lung adenocarcinoma (LUAD) [micro-invasive adenocarcinoma (MIAD), invasive non-mucinous adenocarcinoma, invasive mucinous adenocarcinoma, colloid adenocarcinoma, fetal adenocarcinoma, and entric-type adenocarcinoma], squamous cell carcinomas, adenosquamous cell carcinoma, other types of lung tumors, and metastatic lung cancer. This study excluded squamous cell carcinomas, adenosquamous cell carcinoma, other types of lung tumors, and metastatic lung cancer. The main focus was on lung adenocarcinoma, which was divided into MIAD group and invasive adenocarcinoma (IAD) group (including all types of adenocarcinoma except MIAD). Patients with multiple pulmonary nodules with clear characteristics are grouped based on their pathological results, and the pulmonary nodule with the highest degree of malignancy is included.

### 3. Research Content

#### 3.1 Comparative analysis of clinical characteristics between BSPN and NBSPN

The clinical data and imaging features of patients with BSPN and NBSPN were compared to explore potential high-risk factors for NBSPN.

#### 3.2 Comparative analysis of clinical characteristics between PGL and LUAD

The clinical data and imaging characteristics of patients in the PGL and LUAD were compared to explore the risk factors of LUAD. Additionally, clinical characteristics of MIAD and IAD were compared to explore key factor differentiating between the two.

#### 3.3 Composition of BSPN

The disease composition of BSPN wes analysized to explore reasons for misdiagnosis .

### 4. Statistical Methods

SPSS 27.0 software was used for statistical analysis. Normal distribution measurement data was represented by mean ± standard deviation (SD), independent sample t-test was used for comparison of intergroup differences. Median and interquartile range were used for skewed distribution measurement data, and rank sum test was used for intergroup difference comparison. Count data was represented by frequencies and proportions, and χ ^2^-test was used for intergroup difference comparison. A *p*-value < 0.05 was considered statistically significant. Variables with *P* < 0.1 in univariate logistic regression were considered to be correlated with the research results and further used in stepwise multivariate logistic regression with α = 0.05 as the test criterion, and *P* < 0.05 indicating statistical significance. A clinical prediction model was developed using R-4.4.1 and RStudio 2024.04.2 in the BSPN and NBSPN. The samples were randomly divided into a training set (70%) and a validation set (30%), and a column chart was constructed based on the results of multivariate logistic regression analysis. Receiver Operating Characteristic Curve (ROC) curve of the subjects and calculate the Area Under The Curve (AUC) were drawn to test the performance of our column chart in the training and validation groups. If AUC > 0.75, it was considered that the column chart we established has good predictive performance, while at 0.5-0.75 was considered acceptable.

## Results

### 1. Baseline Clinical Characteristics of Patients

A total of 6,166 patients with pulmonary nodules ≤ 2 cm in diameter were included in this study, of whom 84.5% (5,212/6,166) had undetermined nodule nature, and 15.5% (954/6,166) had confirmed pathology. Among the confirmed cases, 82.1% (783/954) were diagnosed through surgical specimens, 12.9% (123/954) through bronchoscopy biopsy, and 5.0% (48/954) through lung biopsy. Among all patients, 45.4% (2,799/6,166) were male and 54.6% (3,367/6,166) were female. The age distribution was from 9 to 96 years old, with an average age of 53.55 years old. Specifically, 16.0% (986/6,166) were under 40 years old, 51.5% (3,177/6,166) were between 40 and 60 years old, and 32.5% (2,003/6,166) were over 60 years old. Regarding past medical history, the patients with chronic bronchitis accounted for 0.2% (15/6,166), COPD accounted for 0.1% (8/6,166), pulmonary tuberculosis accounted for 0.7% (41/6,166), interstitial pneumonia accounted for 0.0% (1/6,166), and malignant tumor accounted for 4.1% (255/6,166). Additionally, 1.0% (60/6,166) had a history of radiotherapy. A family history of malignant tumors was present in 4.7% (289/6,166), 10.5% (646/6,166) had a smoking history, and 3.8% (232/6,166) had a history of alcohol consumption (Table 1).

**Table 1.** Baseline Characteristics of 6166 Patients with Small Pulmonary Nodules.

|  | Total<br>(n=6166) | Undiagnosed<br>group<br>(n=5212) | Pathologically<br>diagnosed group<br>(n=954) |
| --- | --- | --- | --- |
| Sex (n, %) |  |  |  |
| Female | 3367 (54.6%) | 2795 (53.6%) | 572 (60.0%) |
| Male | 2799 (45.4%) | 2417 (46.4%) | 382 (40.0%) |
| Age (years) | 53.55±13.43 | 53.13±13.73 | 55.85±11.42 |
| <40 (n, %) | 986 (16.0%) | 894 (17.1%) | 92 (9.6%) |
| 40~60 (n, %) | 3177 (51.5%) | 2673 (51.3%) | 504 (52.8%) |
| >60 (n, %) | 2003 (32.5%) | 1645 (31.6%) | 358 (37.5%) |
| Past history (n, %) |  |  |  |
| Chronic bronchitis | 15 (0.2%) | 11 (0.2%) | 4 (0.4%) |
| COPD | 8 (0.1%) | 7 (0.1%) | 1 (0.1%) |
| Pulmonary tuberculosis | 41 (0.7%) | 35 (0.7%) | 6 (0.6%) |
| Interstitial pneumonia | 1 (0.0%) | 1 (0.0%) | 0 (0.0%) |
| Malignant tumor | 255 (4.1%) | 193 (3.7%) | 62 (6.5%) |
| History of radiotherapy<br>(n, %) | 60 (1.0%) | 56 (1.1%) | 4 (0.4%) |
| Family history of malignant<br>tumors (n, %) | 289 (4.1%) | 211 (4.0%) | 78 (8.2%) |
| Smoking history (n, %) | 646 (10.5%) | 508 (9.7%) | 138 (14.5%) |
| Drinking history (n, %) | 232 (3.8%) | 190 (3.6%) | 42 (4.4%) |

In a cohort of the 954 patients with confirmed pathology, 40.0% (382/954) were male and 60.0% (572/954) were female. The age range was 21-82 years old, with an average age of 55.85 years old. Specifically, 9.6% (92/954) were under 40 years old, 52.8% (504/954) were between 40 and 60 years old, and 37.5% (358/954) were over 60 years old. Reviewing their medical history reveals that 0.4% (4/954) had a history of chronic bronchitis, 0.1% (1/954) had a history of COPD, and 0.6% (6/954) had a history of pulmonary tuberculosis. No cases of interstitial pneumonia were reported, and 6.5% (62/954) had experienced malignant tumors. Additionally, 0.4% (4/954) had undergone radiotherapy. A family history of malignant tumors was present in 8.2% (78/954). Smoking and drinking histories were present in 14.5% (138/954) and 4.4% (42 /954) respectively (Table 1).

Among the confirmed cases, 108 (11.3%) were classified as BSPNs, and 846 (88.7%) were categorized as NBSPNs (Figure 2A). Within the NBSPNs, PGL accounted for 14.2% (120/846), with AAH forming a small portion of 5.8% (7/120) and AIS comprising 94.2% (113/120). Lung cancer constituted the largest share of 85.8% (726/846), with adenocarcinoma comprising 87.2% (633/726), squamous cell carcinoma comprising 2.9% (21/726), adenosquamous cell carcinoma comprising 0.4% (3/726), other types of lung tumors comprising 4.1% (30/726), and metastatic lung cancer comprising 5.4% (39/726). Among the LUAD, MIAD represented 32.1% (203/633), and IAD accounted for 67.9% (430/633). In the 108 benign nodule cases, non-specific inflammatory nodules (NSIN) were most common, accounting for 57.4% (62/108), followed by lung specific granulomatous nodules at 9.3% (10/108), fungal infections at 11.1% (12/108), hamartomas at 8.3% (9/108), lymph nodes at 1.9% (2/108), and others contributing to the remaining 12.0% (13/108) (Figure 2B, Table S1 in the Appendix).

**Figure 2.**
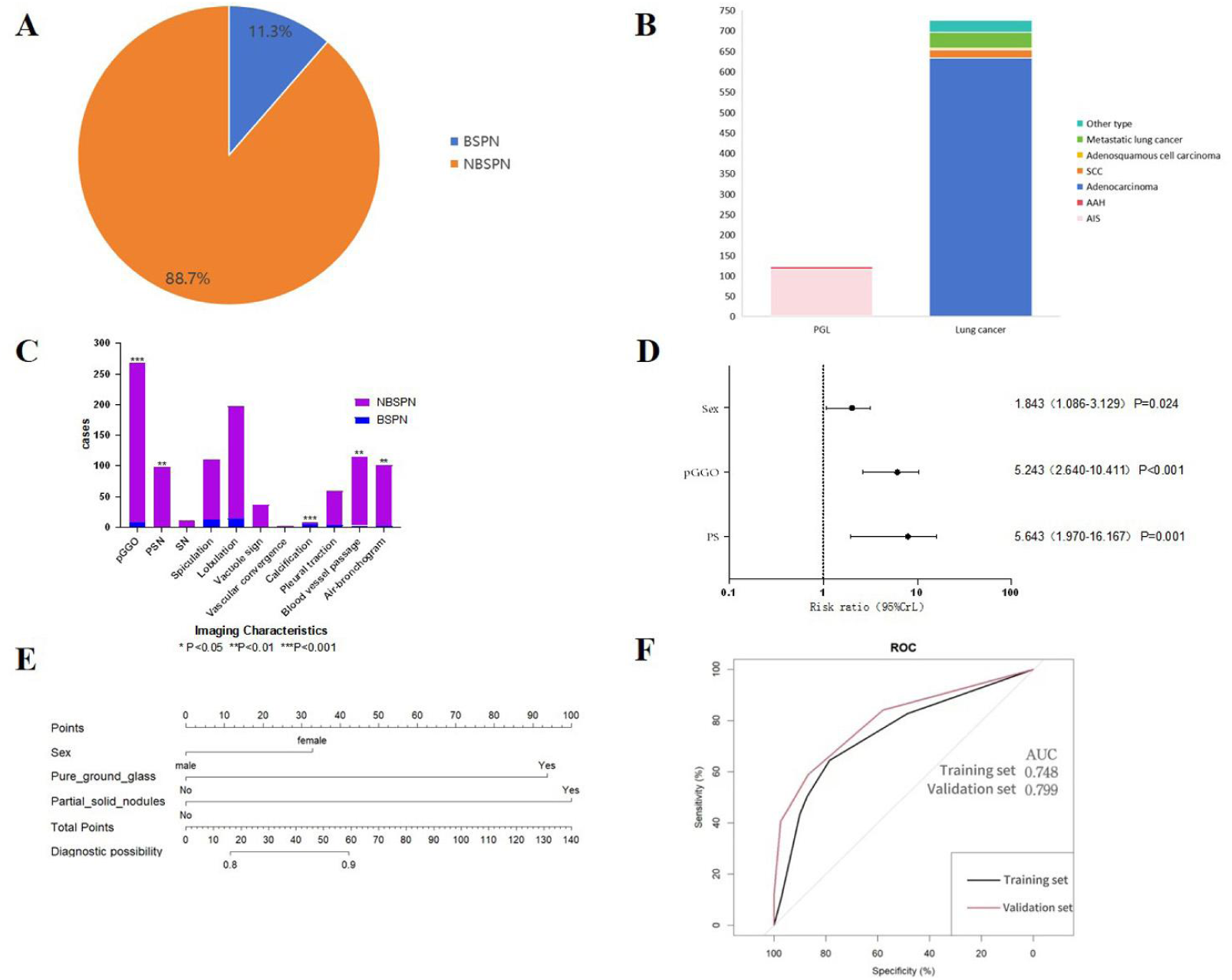
(A) The proportion of BSPN and NBSPNsin pathologically confirmed SPN. (B) Composition of the Nature of SPN in 954 Patients. (C) Comparison of Imaging Characteristics Between BSPN and NBSPN (\**P* < 0.05, \*\**P* < 0.01, \*\*\**P* < 0.001). (D) Forest Plot of Multifactorial Logistic Regression Analysis of Imaging Characteristics Between BSPN and NBSPN. (E) Nomogram for Predicting the risk of NBSPN (Scores from variables like Sex, pGGO, and PSN are summed to find the Total Points, which corresponds to the Diagnostic Probability of the nodule being non-benign.). (F) ROC Curve Analysis of Training Set and Validation Set Nomograms. AUC is reported. Solid line represents the ROC curve. Dotted diagonal line represents the line of equality.

### 2. Comparison of Characteristics Between BSPN and NBSPN

#### 2.1 Comparison of Clinical Data Between BSPN and NBSPN

Analysis of clinical data highlighted a statistically significant difference between BSPN and NBSPN in terms of gender (*P* < 0.001). Conversely, no significant differences were observed in age, history of malignant tumors, family history of malignant tumors, smoking history, or levels of tumor markers (CEA, CYFRA21-1, SCC, NSE, proGRP) (*P* > 0.05) (Table 2).

**Table 2.**
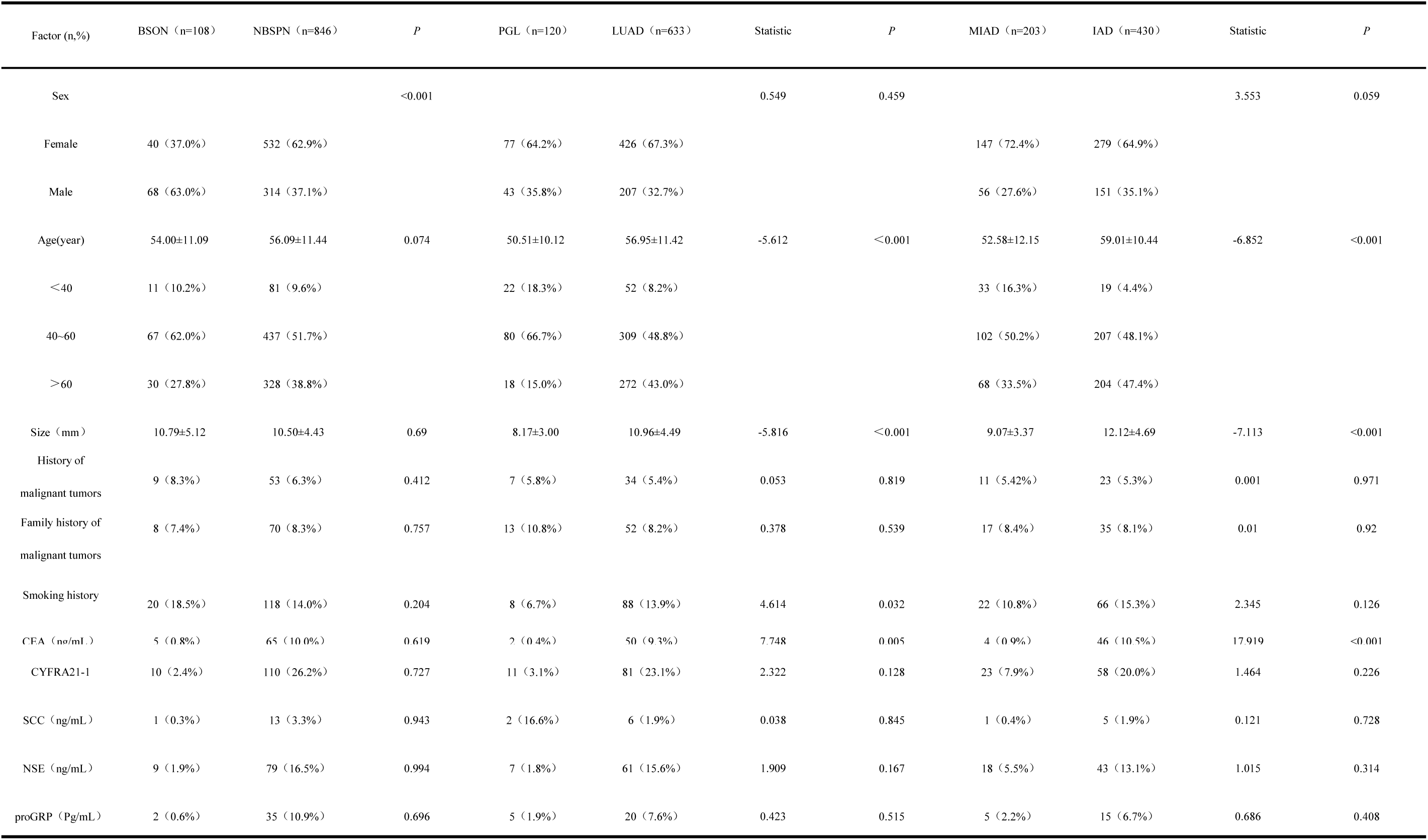
Comparison of Clinical Data Between BSPN and NBSPN, PGL and LUAD, MIAD and IAD.

#### 2.2 Comparison of Size and Imaging Features Between BSPN and NBSPN

Among the 954 patients with confirmed nodule characteristic and complete imaging data, 108 cases were identified as BSPNs (11.3%), and 846 were categorized as NBSPNs (88.7%).

Through the analysis of basic clinical data from two groups of patients, it was found that there were statistically significant differences in several imaging features of nodules, including pGGO (*P* < 0.001), PSN (*P* = 0.003), calcification (*P* < 0.001), blood vessel passage (*P* = 0.009), and air-bronchogram (*P* = 0.007), while there were no statistically significant differences in SN, spiculation, spinous processes, vacuoles, vasculore sign and pleural traction (*P* > 0.05) (Figure 2C, Table S2 in the appendix).

The 954 patients were randomly divided into a training set (n = 670) and a validation set (n = 284) at a ratio of 7:3. Logistic regression analysis was performed on the training set’s clinical features. Factors with *P* < 0.05 in multivariate analysis were included in the clinical prediction model. These factors encompassed gender, pGGO appearance, and PSN appearance (Figure 2C, D). These features were incorporated into a nomogram (Figure 2E), which illustrated the relative contribution of each factor to the probability of a nodule being non-benign (the higher the risk factor score, the greater the probability of being non-benign). ROC curves were generated to assess the stability of the nomogram. AUC values for the training and validation sets were 0.748 and 0.799 (Figure 2F).

### 3. Comparison of Clinical Characteristics Between PGL and LUAD

#### 3.1 Comparison of Clinical Data Between PGL and LUAD

Excluding cases of squamous cell carcinoma, adenosquamous cell carcinoma, other types of lung tumors, and metastatic lung cancer, a total of 93 cases were excluded, remaining 753 cases in the lung cancer group. Among these, 120 cases [15.9% (120/753)] were identified as PGL, and 633 cases [84.1% (633/753)] were classified as LUAD. Comparison of clinical data revealed statistically significant differences between the two groups in terms of age (*P* < 0.001), smoking history (*P* = 0.032), and CEA levels (*P* = 0.005). No significant differences were observed regarding gender, history of malignant tumors, family history of malignant tumors, or levels of CYFRA21-1, SCC, NSE, and proGRP (*P* > 0.05) (Table 2).

#### 3.2 Comparison of Size and Imaging Features Between PGL and LUAD

Within the lung cancer group, a total of 513 nodules with clear characteristics and complete imaging data were analyzed. The mean size of nodules in the PGL was 8.17 ± 3.00 mm, signigicantly smaller than the 10.96 ± 4.49 mm observed in the LUAD (*P* < 0.001). Signigicant differences in imaging features were noted between the two groups in terms of pGGO (*P* < 0.001), PSN (*P* = 0.004), spiculation (*P* < 0.001), lobulation (*P* = 0.001), and pleural traction (*P* = 0.025). No significant differences were observed in SN, spinous processes, vacuole sign, vascular convergence, calcification, blood vessel passage, or air-bronchogram (*P* > 0.05) (Figure 3A, Table S3 in the Appendix).

**Figure 3.**
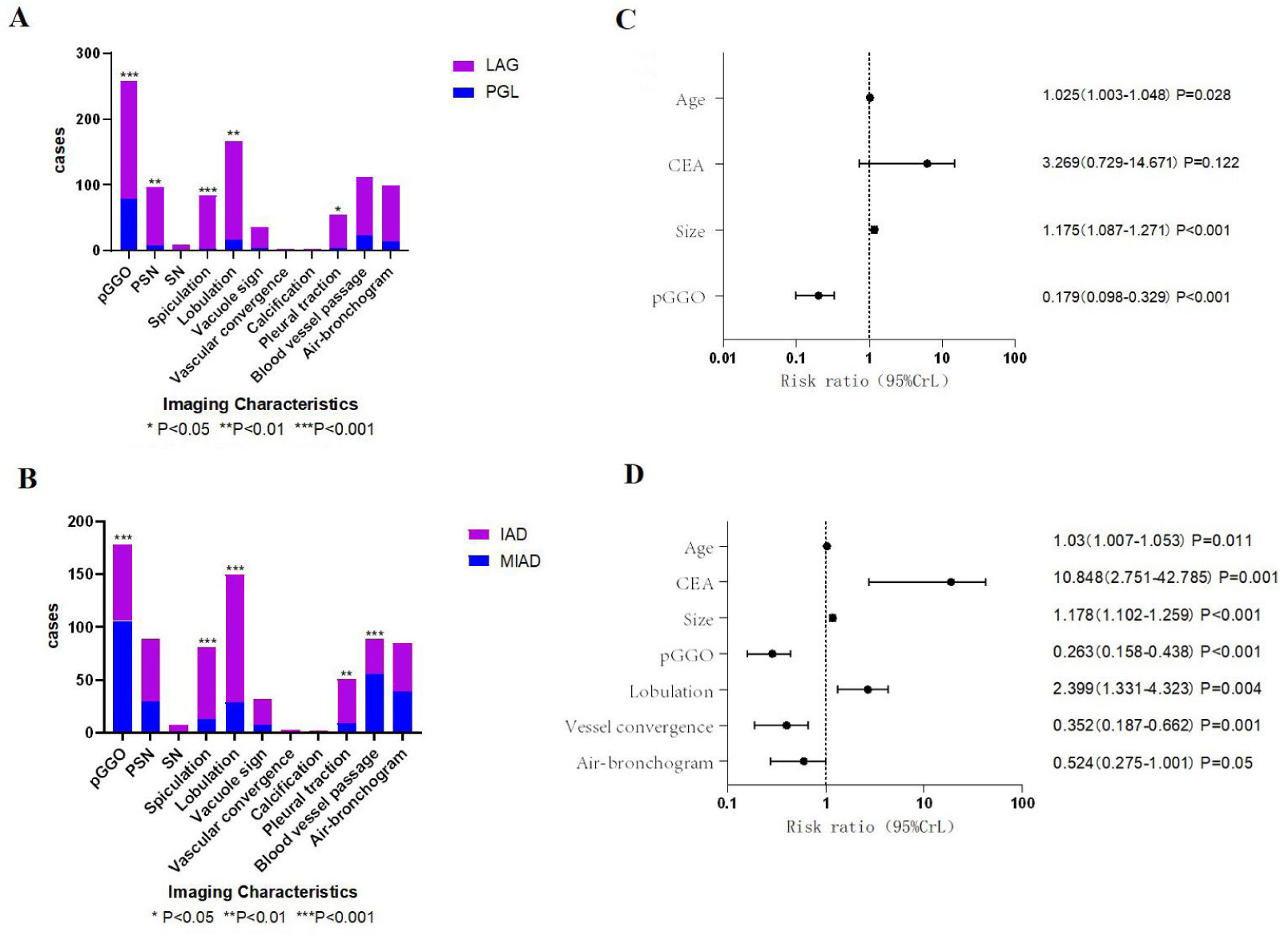
(A) Comparison of Imaging Characteristics Between PGL and LUAD. (B) Forest Plot of Multifactorial Logistic Regression Analysis of Imaging Characteristics Between PGL and LUAD. (C) Comparison of Size and Imaging Characteristics Between MIAD and IAD. (D) Forest Plot of Multifactorial Logistic Regression Analysis of Imaging Characteristics Between MIAD and IAD.

Univariate logistic regression analysis of clinical characteristics in the PGL and LUAD revealed that age (*P* = 0.028) and nodule size (*P* < 0.001) were positively correlated with LUAD, whereas pGGO (*P* < 0.001) showed a negative correlation (Table 3, Figure 3B).

**Table 3.**
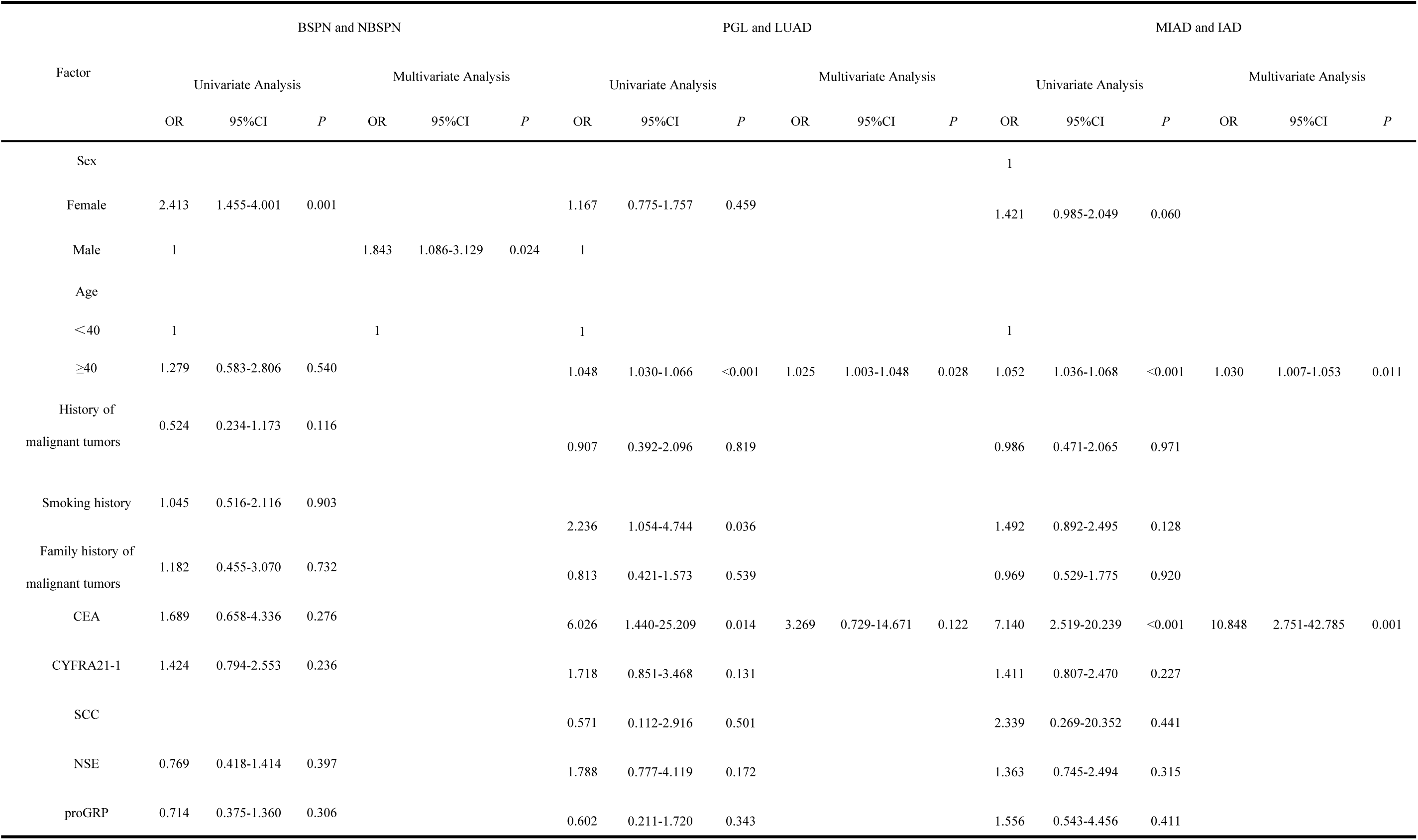
Logistic Regression Analysis of Clinical Characteristics for the Training Groups in BSPN and NBSPN, PGL and LUAD, MIAD and IAD.

#### 3.3 Comparison of Clinical Characteristics Between MIAD and IAD

##### 3.3.1 Comparison of Clinical Data Between MIAD and IAD

Among the 633 cases of LUAD, MIAD accounted for 32.1% (203/633), and IAD constituted 67.9% (430/633). Analysis revealed statistically significant differences between the two groups in age (*P* < 0.001) and CEA levels (*P* < 0.001). No significant differences were observed in gender, history of malignant tumors, family history of malignant tumors, smoking history, or levels of CYFRA21-1, SCC, NSE, and proGRP (*P* > 0.05) (Table 2).

##### 3.3.2 Comparison of Size and Imaging Features Between MIAD and IAD

The average size of nodules in the MIAD was 9.07 ± 3.37 mm, compared to 12.12 ± 4.69 mm for IAD, demonstrating a statistically significant difference (*P* < 0.001) (Table 2). Significant differences were also observed in imaging features such as pGGO (*P* < 0.001), spiculation (*P* < 0.001), lobulation (*P* < 0.001), pleural traction (*P* = 0.002), and vascular traversal (*P* < 0.001). Conversely, no significant differences were observed in PSN, SN, spinous processes, vacuole sign, vascular convergence, calcification, or air-bronchogram (*P* > 0.05) (Figure 3C, Table S4 in the Appendix).

Univariate logistic regression analysis of clinical characteristics in the MIAD and IAD revealed that age (*P* = 0.011), CEA levels (*P* = 0.001), nodule size (*P* < 0.001), and lobulation (*P* = 0.004) were positively correlated with IAD. In contrast, pGGO (*P* < 0.001), blood vessel passage (*P* = 0.001), and air-bronchogram (*P* = 0.050) were negatively correlated (Table 3, Figure 3D).

## Discussion

Lung cancer stands as the foremost cause of cancer-related mortality worldwide and ranks first among malignant tumors in terms of incidence and mortality in China. With the implementation of the Healthy China strategy, the widespread adoption of national health examinations and screenings, along with continuous advancement in medical technology, the detection rate of pulmonary nodules, especially those ≤ 2 cm in diameter, has been on an upward trend during routine health check-ups each year. A multicenter lung cancer screening study conducted in China showed that low-dose computed tomography (LDCT) detected pulmonary nodules in 29.89% of the screened population,[12] with the majority measuring less than 2 cm in diameter. This has attracted widespread attention. Clinically, lung cancer is often diagnosed at an advanced stage, leaving only 15% of patients eligible for surgical intervention, and resulting in a 5-year survival rate of merely 15%. In contrast, the early stage lung cancer rate detected by screening for pulmonary nodules is as high as 80%, with 80% to 90% of patients achieving nearly a 100% 5-year survival rate following surgery. Therefore, distinguishing between benign and malignant pulmonary nodules, especially those with a diameter of ≤ 2 cm, holds substantial clinical significance for selecting appropriate treatments and developing reasonable screening and follow-up strategies. At present, most experts agree that pulmonary nodules are predominantly focus on pulmonary nodules with a diameter of ≤ 3 cm.[13] There are few reports on the research of pulmonary nodules with a diameter of ≤ 2 cm, and there is a scarcity of studies analyzing and comparing the characteristics of different types of pulmonary nodules such as PGL, MIAD, and IAD. Consequently, this study intends to use real-world data to analyze the disease spectrum of pulmonary nodules with a diameter of ≤ 2 cm, identify the risk factors for NBSPN, and characterize different types of nodules. The ultimate goal is to provide critical references for improving clinical accuracy in discrimination and decision-making.

In this study, a total of 6,166 cases were followed up, with 15.5% (954/6,166) having confirmed pathology for pulmonary nodules ≤ 2 cm in diameter. NBSPN accounted for 13.7% (846/6,166), and LUAD cases comprised 10.3% (633/6,166) of the total population, figures that are higher than those reported in other studies.[14] This discrepancy may be related to the inclusion criteria of this study, which focused on patients who had undergone at least one follow-up visit at the hospital.

This study found that NBSPNs were more prevalent in female patients compared to BSPNs. However, no significant differences were observed in age, smoking history, the history of malignant tumors, or family history of malignant tumors between the two groups. This observation contrasts with several studies and established guidelines,[15][16][17] including the National Comprehensive Cancer Network (NCCN) guidelines for non-small cell lung cancer (NSCLC) and the Chinese guidelines for LDCT screening of lung cancer,[18] which identify age, smoking history,[19] previous malignant tumor history,[20] and family history of lung cancer[21][22] as high-risk factors for lung cancer. Interestingly, this study did not find any differences in smoking history and family history of lung cancer between the two groups, which is consistent with some studies.[19][23][24] This may be related to the higher proportion of female patients and non-smokers in the population included in this study. However, this study found that smoking history and family history of lung cancer were not significantly correlated with NBSPN, suggesting a potential decrease in the relevance of these factors in the malignant transformation of small nodules. Alternatively, it might reflect the impact of industrialization leading to environmental pollution, thereby increasing the incidence of gene mutations and enhancing the risk of malignant transformation of small nodules.[23][25][26]

For patients with pulmonary nodules ≤ 2 cm in diameter, typical imaging changes are often not prominent. However, this study revealed that pGGO appearance of pulmonary nodules is associated with a higher probability of NBSPN, while calcification indicates a higher likelihood of BSPN, consistent with previous clinical studies.[27–30] Given the limitations of using only two imaging features to determine nodule nature, a clinical prediction model was developed to predict the probability of benign or malignant pulmonary nodules. Gender, pGGO, and PSN were included as features in the nomogram. The study demonstrated that PSN appearance serves as the strongest predictor of NBSPN, followed by pGGO.

This article further compares LUAD and PGL in NBSPN, revealed that age is associated with the likelihood of developing LUAD. Specifically, the older the patient, the greater the possibility of LUAD. In this study, 91.8% of patients diagnosed with LUAD were over 40 years old, supporting the definition of high-risk population for lung cancer in the 2024 Chinese Expert Consensus on the Diagnosis and Treatment of Pulmonary Nodules.[8] The lack of statistical significance in serum CEA levels may be attributed to the relatively low tumor burden and limited secretion of tumor markers into the bloodstream. Additionally, the study found that pGGO is more likely associated with PGL, aligning with previous studies.[31][32] Furthermore, the size of nodules plays a crucial role in evaluating whether pulmonary nodules are infiltrative growth. The average size of PGL was 8.17 mm, while that of LUAD was 10.96 mm, indicating a positive correlation between nodule size and malignancy risk. Therefore, in clinical practice, nodules approximately 8 mm in size with pGGO appearance are more likely to be PGL. This finding supports the viewpoint mentioned in the 2024 Chinese Expert Consensus on the Diagnosis and Treatment of Pulmonary Nodules,[8] which states that for nodules with a diameter greater than 8 mm, the probability of malignancy of pulmonary nodules can be evaluated qualitatively using clinical judgment and/or quantitatively using validation models. The proliferation rate of the PGL is slower, and close observation can be given. If there is a trend of tumor proliferation such as nodule enlargement or density increase, it is a better surgical intervention timing. However, the specific timing of surgical intervention still needs to be further explored.

This article further explored the clinical differences between MIAD and IAD within LUAD. It was found that the older age and the higher the serum CEA level are associated with an increased likelihood of IAD. However, it is noteworthy that the CEA elevation rate was very low, accounting for only 10.5%. Therefore, imaging features and necessary follow-up examinations remain crucial for accurate diagnosis. In terms of imaging features, this study revealed that the probability of nodules being MIAD increased when the nodules appeared as pGGO and blood vessel passage. Conversely, the presence of lobulation increase the possibility of a nodule being IAD, which is consistent with some clinical studies.[23][33] In addition, in this study, the average size of the MIAD was 9.07 mm, while IAD averaged 12.12 mm. The larger the nodule, the more likely it is to be IAD, which may be related to the faster growth rate of tumors with higher malignancy. This finding differs from some clinical studies,[27] suggesting that nodule size can also serve as an important reference for distinguishing MIAD from IAD.

In this study, some postoperative pathological results indicated BSPN, with non-specific inflammatory nodules being the most common type,[34][35] accounting for 57.4% (62/108). Other types of BSPNs included NSG, fungus, hamartoma, and lymph nodes. BSPNs accounted for 11.3% (108/954) of all confirmed pulmonary nodules in this cohort. Therefore, over 10% of patients with pulmonary nodules underwent unnecessary surgical treatment due to misdiagnosis as NBSPNs. This highlights the critical need for clinicians to conduct a more thorough analysis of clinical features before opting for surgery to accurately assess the probability of benign or malignant pulmonary nodules, thereby avoiding unnecessary invasive surgeries.

In summary, this study found that among patients with pulmonary nodules ≤ 2 cm in diameter who were followed up, approximately 13% were NBSPNs. When there are no obvious clinical symptoms and the imaging shows calcification, the probability of BSPN is higher. In contrast, a pGGO appearance on imaging should raise suspicion for NBSPNs. Besides, when assessing small pulmonary nodules (SPNs), attention should be given to the following indicators to identify potential NBSPNs: gender (female), imaging features (pGGO or PSN), with PSN being the strongest risk factor. Additionally, careful evaluation of nodule size is crucial. If the nodule diameter exceeds 8 mm but does not exceed 10 mm, there is a higher likely of it being PGL. If the nodule size is greater than 10 mm and the patient is over 40 years old, the possibility of LUAD should be highly considered. A higher possibility of IAD if serum CEA levels are elevated. However, the rate of CEA elevation is low, and even with normal CEA levels, the possibility of IAD cannot be completely ruled out. Therefore, close follow-up is still necessary.

This study has certain limitations: 1. This was a retrospective study, and data were retrieved from a clinical research big data platform. Some clinical data may be missing or incomplete, A large-scale prospective study is needed to further validate these findings. 2. The data were from a single center and need to be verified through multicenter studies. 3. The follow-up data for pulmonary nodules were not rigorous or standardized, making it difficult to conduct correlation studies on the differentiation of benign and malignant nodules. Future research should focus on multicenter prospective studies to address these limitations.

## Ethical approval

All patients signed informed consent and the ethics approval was granted by the Ethics Committee of Fujian Provincial Hospital (No. K-2025-02-153).

## Data Availability

All data produced in the present study are available upon reasonable request to the authors.

