## supplement table for "Differential Diagnosis of Image Discovered Small Pulmonary Nodules (SPN): A Real-World Study": Table S1.docx

Table S1. Composition of the Nature of Small Pulmonary Nodules in 954 Patients

| Nature of Small Pulmonary Nodules |  |
| --- | --- |
| Benign（n，%） | 108（11.3%） |
| NSIN（n，%） | 62（57.4%） |
| NSG（n，%） | 10（9.3%） |
| Fungus（n，%） | 12（11.1%） |
| Hamartoma（n，%） | 9（8.3%） |
| Lymph node（n，%） | 2（1.9%） |
| Other（n，%） | 13（12.0%） |
| Non-benign（n，%） | 846（88.7%） |
| PGL（n，%） | 120（14.2%） |
| AAH（n，%） | 7（5.8%） |
| AIS（n，%） | 113（94.2%） |
| Lung cancer（n，%） | 726（85.8%） |
| LUAD | 633（87.2%） |
| MIAD（n，%） | 203（32.1%） |
| IAD（n，%） | 430（67.9%） |
| SCC（n，%） | 21（2.9%） |
| AC（n，%） | 3（0.4%） |
| Other（n，%） | 30（4.1%） |
| MC（n，%） | 39（5.4%） |
