## supplement table for "Differential Diagnosis of Image Discovered Small Pulmonary Nodules (SPN): A Real-World Study": Table S2.docx

Table S2. Comparison of Size and Imaging Characteristics Between Benign and Non-benign Small Pulmonary Nodule Groups

|  | BSPN（n=108） | NBSPN（n=846） | t | *P* |
| --- | --- | --- | --- | --- |
| pGGO（n，%） | 8（7.4%） | 260（30.7%） | 20.323 | ＜0.001 |
| PS（n，%） | 1（0.9%） | 97（11.5%） | 8.896 | 0.003 |
| SN（n，%） | 1（0.9%） | 10（1.2%） | 0.001 | 1.000 |
| Spiculation（n，%） | 13（12.0%） | 97（11.5%） | 1.421 | 0.233 |
| Spinous process（n，%） | 0（0.0%） | 0（0.0%） |  |  |
| Lobulation（n，%） | 14（13.0%） | 184（21.7%） | 1.184 | 0.277 |
| Vacuole sign（n，%） | 1（0.9%） | 36（4.3%） | 1.858 | 0.287 |
| Vessel convergence（n，%） | 0（0.0%） | 3（0.4%） | 0.294 | 1.000 |
| Calcification（n，%） | 5（4.6%） | 3（0.4%） | 28.777 | ＜0.001 |
| Pleuralre traction（n，%） | 4（3.7%） | 56（6.6%） | 0.404 | 0.525 |
| Blood vessel passage（n，%） | 3（2.8%） | 112（13.2%） | 6.873 | 0.009 |
| Air-bronchogram（n，%） | 2（1.9%） | 100（11.8%） | 7.233 | 0.007 |
