## supplement table for "Differential Diagnosis of Image Discovered Small Pulmonary Nodules (SPN): A Real-World Study": Table S3.docx

Table S3. Comparison of Size and Imaging Characteristics Between the Precursor Glandular Lesions Group and the Lung Adenocarcinoma Group

|  | PGL  （n=120） | LUAD（n=633） | t | *P* |
| --- | --- | --- | --- | --- |
| pGGO（n，%） | 79（84.0%） | 179（42.7%） | 52.439 | ＜0.001 |
| PS（n，%） | 8（8.5%） | 89（21.2%） | 8.115 | 0.004 |
| SN（n，%） | 1（1.1%） | 8（1.9%） | 0.318 | 0.897 |
| Spiculation（n，%） | 3（3.2%） | 81（19.3%） | 14.606 | ＜0.001 |
| Spinous process（n，%） | 0（0.0%） | 0（0.0%） |  |  |
| Lobulation（n，%） | 17（18.1） | 150（35.8%） | 10.973 | 0.001 |
| Vacuole sign（n，%） | 4（4.3%） | 32（7.6%） | 1.346 | 0.246 |
| Vessel convergence（n，%） | 0（0.0%） | 3（0.7%） | 0.677 | 1.000 |
| Calcification（n，%） | 1（1.1%） | 2（0.5%） | 0.454 | 0.456 |
| Pleuralre traction（n，%） | 4（4.3%） | 51（12.2%） | 5.027 | 0.025 |
| Blood vessel passage（n，%） | 23（24.5%） | 89（21.2%） | 0.468 | 0.494 |
| Air-bronchogram（n，%） | 15（16.0%） | 85（20.3%） | 0.917 | 0.338 |
