## supplement table for "Differential Diagnosis of Image Discovered Small Pulmonary Nodules (SPN): A Real-World Study": Table S4.docx

Table S4. Comparison of Size and Imaging Characteristics Between the Microinvasive Adenocarcinoma Group and the Invasive Adenocarcinoma Group

|  | MIAD  （n=203） | IAD  （n=430） | t | *P* |
| --- | --- | --- | --- | --- |
| pGGO（n，%） | 106（52.2%） | 72（16.7%） | 63.926 | ＜0.001 |
| PS（n，%） | 30（14.8%） | 59（13.7%） | 0.715 | 0.398 |
| SN（n，%） | 1（0.5%） | 7（1.6%） | 2.184 | 0.139 |
| Spiculation（n，%） | 13（6.4%） | 68（15.8%） | 19.822 | <0.001 |
| Spinous process（n，%） | 0（0.0%） | 0（0.0%） |  |  |
| Lobulation（n，%） | 29（14.3%） | 121（28.1%） | 33.140 | <0.001 |
| Vacuole sign（n，%） | 8（3.9%） | 24（5.6%） | 2.331 | 0.127 |
| Vessel convergence（n，%） | 0（0.0%） | 3（0.7%） | 1.818 | 0.178 |
| Calcification（n，%） | 1（0.5%） | 1（0.2%） | 0.133 | 0.716 |
| Pleuralre traction（n，%） | 9（4.4%） | 42（9.8%） | 9.821 | 0.002 |
| Blood vessel passage（n，%） | 56（27.6%） | 33（7.7%） | 31.012 | <0.001 |
| Air-bronchogram（n，%） | 39（19.2%） | 46（10.7%） | 3.151 | 0.076 |
